# Novel echocardiographic markers of elevated left ventricular filling pressure during diastolic stress testing

**DOI:** 10.1101/2020.12.16.20248175

**Authors:** Jan Verwerft, Frederik H. Verbrugge, Guido Claessen, Lieven Herbots, Paul Dendale, Andreas B. Gevaert

**Author notes:** Corresponding author: Andreas B. Gevaert, Research Group Cardiovascular Diseases, Department GENCOR, University of Antwerp, Universiteitsplein 1, 2610 Antwerp, Belgium.

## Abstract

**Aims:** Diastolic stress testing (DST) is recommended to confirm heart failure with preserved ejection fraction (HFpEF) in patients with exertional dyspnea, but algorithms lack sensitivity. We aimed to identify additional echocardiographic markers of elevated pulmonary arterial wedge pressure during exercise (exPAWP) in patients referred for DST.

**Methods and Results:** We analyzed 22 patients referred for exercise right heart catheterization with simultaneous echocardiography. We identified candidate parameters in patients with exPAWP ≥25 mmHg. Elevated exPAWP was present in 14 patients, and was best identified by peak septal systolic annular velocity on color Doppler (*exS*’, area under the receiver operating characteristic curve (AUC) 0.97, 95% confidence interval 0.92-1.0) and mean pulmonary artery pressure/cardiac output slope (mPAP/CO, AUC 0.88 [0.72-1.0]). We propose a three-step decision tree to identify patients with elevated exPAWP. Applying this decision tree to 376 patients in an independent non-invasive DST cohort showed that patients labeled as ‘high probability of HFpEF’ had reduced peak oxygen uptake (12.8 (10.5-15.9) mL/kg/min, p<.001 vs intermediate/low probability), high H2FPEF score (55 (44-75)%, p<.007 vs intermediate/low probability), and typical clinical characteristics. The amount of inconclusive DST decreased from 80% using current recommendations, to 29% using the decision tree.

**Conclusion:** In DST for suspected HFpEF, *exS*’ was the most accurate echocardiographic parameter to identify elevated PAWP. We propose a decision tree including *exS*’ and mPAP/CO for interpretation of DST. Application of this decision tree revealed typical HFpEF characteristics in patients labeled as high probability of HFpEF, and substantially reduced the amount of inconclusive results.

## Introduction

Half of heart failure (HF) patients have a preserved ejection fraction (HFpEF) ^1^. Compared to HF with reduced ejection fraction, the diagnosis of HFpEF is often more challenging, especially when patients are not decompensated ^2^. Guidelines recommend using the combination of patient characteristics, natriuretic peptide levels, and echocardiography at rest to make a diagnosis of HFpEF ^3,4^. However, in patients without gross volume overload who complain from chronic dyspnea, a diagnosis of HFpEF can be easily missed at rest, as many patients only develop symptoms and disproportionate elevation of cardiac filling pressures during exercise ^5^.

Invasive hemodynamic exercise testing is considered the gold standard to rule in or rule out HFpEF based on a pulmonary arterial wedge pressure (PAWP) ≥25 mmHg or <25 mmHg during symptom-limited supine exercise (exPAWP) ^6^. Yet, this strategy is not broadly applied due to logistic restraints and limited expertise. A positive diastolic stress test (DST) in patients with an intermediate to high pretest probability may offer a valuable alternative to confirm the diagnosis of HFpEF, with this approach supported by a recent consensus statement of the Heart Failure Association of the European Society of Cardiology ^7^. DST refers to the use of echocardiography to detect impaired left ventricular (LV) diastolic functional reserve and disproportionally increased filling pressures during exercise that commonly result in pulmonary hypertension ^8^. Accordingly, elevated early mitral inflow velocity over early diastolic annular velocity (*E/e*’) and tricuspid regurgitation (TR) velocity during exercise (*exE/e*’, exTR) are used to support a diagnosis of HFpEF ^9,10^. Although the positive predictive value of the DST is acceptable at 85-93%, its negative predictive value is poor (55-77%) ^11^.

In this study, the aim was to identify additional echocardiographic markers of elevated PAWP ≥25 mmHg assessed by gold-standard invasive hemodynamic exercise testing performed because of unexplained exertional dyspnea. Subsequently, we aimed to apply these echocardiographic parameters in an independent cohort referred for non-invasive DST.

## Methods

### Study population

We performed a retrospective analysis of patients referred to Jessa Hospital (Hasselt, Belgium) because of exertional dyspnea not sufficiently explained by resting examinations. We screened patients referred from April 2017 to May 2020. Inclusion criteria were: left ventricular ejection fraction (LVEF) ≥50%, no more than mild valvular stenosis at rest, no more than moderate left-sided valvular insufficiency at rest, and no indication that pulmonary disease was the sole cause of exertional dyspnea (as assessed by the referring physician). Non-invasive DST was performed in all consecutive patients (DST cohort). If non-invasive DST was inconclusive, patients were offered invasive hemodynamic exercise testing with simultaneous echocardiography and gas exchange measurement (exRHC cohort). We used the exRHC cohort for derivation of the echocardiographic variables associated with elevated PAWP. We applied these novel variables to the DST cohort. Patients included in the exRHC cohort were excluded from validation analyses in the DST cohort. This study complies with the Declaration of Helsinki and was approved by the Ethical Committee of the Jessa Hospital. All patients provided informed consent.

### Study protocol

All patients underwent a cardiopulmonary exercise test (CPET) with respiratory gas analysis (CS-200, Schiller). Exercise was performed on a semi-supine bicycle ergometer (ErgoLine) with a continuous ramp protocol aimed for a total exercise duration of 10-12 min. In the DST cohort, 2 stage holds were performed at the aerobic threshold and at peak exercise for image acquirement. In the exRHC cohort, a pulmonary artery catheter (Edwards Lifesciences) was placed under fluoroscopic guidance at the catheterization lab before start of the CPET. In addition, the right radial artery was cannulated with a 5F arterial catheter. The fluid filled catheters were then connected to a pressure transducer unit (PowerLab, ADInstruments) with zeroing at the mid axillary level. Every 3 minutes during exercise and at peak exercise, arterial and mixed venous blood gas samples were obtained and PAWP was measured. Other hemodynamic measurements were registered continuously. Echocardiography data was simultaneously collected at aerobic threshold and peak exercise. Hemodynamic tracings were stored in LabChart v8.1 (ADInstruments) for offline analysis by an experienced cardiologist blinded to echocardiographic measurements (J.V.). All pressure measurements were performed at end-expiration by averaging at least 3 cardiac cycles. Cardiac output (CO) was calculated using the Fick method.

### CPET measurements

Ventilation (VE), oxygen uptake (VO_2_) and carbon dioxide production (VCO_2_) were continuously measured through a face mask during exercise. The aerobic threshold was defined as a sustained rise in O_2_ ventilatory equivalent, the anaerobic threshold was defined as a sustained rise in CO_2_ ventilatory equivalent. Peak VO_2_ was defined as the highest 10-second average of VO_2_ during exercise ^12^.

### Echocardiographic measurements

Measurements were performed offline using EchoPAC software (GE Healthcare) according to current guidelines ^10,13^. Examinations were performed with Vivid E9 (GE Healthcare). Peak mitral systolic annular velocity (*S’*) was assessed on color tissue Doppler imaging (TDI) at the level of the septal mitral annulus (Supplemental Figure 1, Supplemental File 1). Medial *e*’ was measured at the septal mitral valve annulus using pulse wave TDI. Systolic pulmonary artery pressure (sPAP) was estimated from TR velocity without adding right atrial pressure. Colloid enhancement of the tricuspid insufficiency signal was systematically employed as previously described ^14^. Mean PAP (mPAP) was calculated by the Chemla formula as sPAP*0.61+2. Stroke volume (SV) and CO were calculated using the left ventricular outflow tract method.

**Figure 1:**
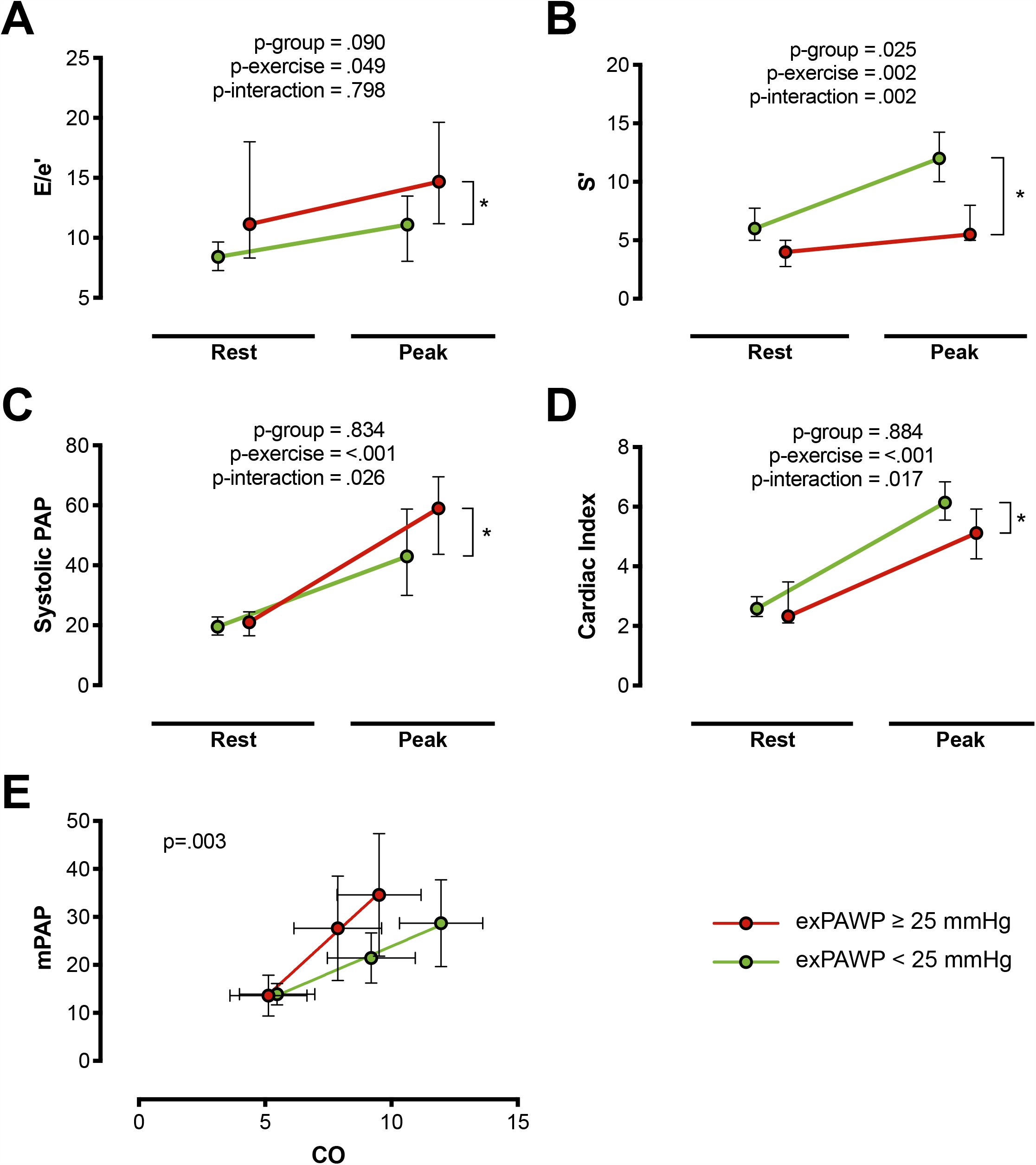
DST parameters associated with elevated exPAWP. Results of non-invasive E/e’ (A), S’ (B), systolic PAP (C), cardiac index (D), and mean PAP/CO slope (E) in the exRHC cohort at rest and peak exercise. Red: patients with elevated exPAWP (n=14), green: patients with normal exPAWP (n=8). P values from linear mixed models. * p <.05 in multiplicity-adjusted comparison of peak values. CO = cardiac output, exPAWP = pulmonary artery wedge pressure during peak exercise, mPAP = mean pulmonary artery pressure, PAP = pulmonary artery pressure.

In 22 patients, measurements were repeated twice in a blinded fashion by 3 observers, to assess intra-observer variability.

### Definitions and thresholds

Elevated cardiac filling pressures were primarily defined as a peak exercise PAWP ≥25 mmHg on invasive hemodynamic assessment, and alternatively as PAWP/CO slope ≥2.0 mmHg/L ^5,15^. Exercise pulmonary hypertension was defined as mPAP/CO slope ≥3.0 mmHg/L by invasive hemodynamic assessment, and ≥3.2 mmHg/L by echocardiography, as previous studies reported higher values on echocardiography ^14^.

Rule-in and rule-out of HFpEF on non-invasive DST was defined according to the most recent American Society of Echocardiography (ASE) and European Association of Cardiovascular Imaging (EACVI) recommendations ^9,10^. HFpEF was diagnosed when septal *exE/e’* ≥15, exTR >2.8 m/s and baseline *e’* <7 cm/s, HFpEF was ruled out when septal *exE/e’* <10 and exTR <2.8 m/s ^9,10^.

To evaluate the performance of the novel echocardiographic markers of elevated exPAWP, the probability of HFpEF according to the novel marker was compared to surrogate HFpEF indicators: peak VO_2_ and logistic H2FPEF score. The latter calculates the probability of HFpEF through clinical and echocardiographic parameters, and has been developed using invasive exRHC measurements ^16^.

### Sample size calculation

Using f test power calculation for repeated measures (GPower v3.1.9), we estimated that a sample of 16 patients would provide 90% power to detect a difference in echocardiographic parameters between elevated and normal PAWP. Effect size was based on the difference in *exE/e’* in the study by Obokata et al ^11^.

### Statistical analysis

Continuous variables were expressed as mean ± standard deviation or median (interquartile range) in case of a skewed distribution. Categorical variables were expressed as percentages. Baseline comparisons were performed using Mann-Whitney-U test, Pearson’s Chi-squared test or Fisher’s Exact test where appropriate. Comparisons between 3 groups were performed using Kruskal-Wallis test with Dunn test for between-group comparisons (continuous variables), and Pearson’s Chi-squared test with pairwise nominal independence test (categorical variables). Interobserver variability was calculated using a two-way agreement intra-class correlation model and using Bland-Altman plots.

DST parameters were compared between patients with elevated vs. normal exPAWP using Mann-Whitney-U test (single measurement during DST, for example mPAP/CO slope) or linear mixed models (repeated measurement during DST, for example *E/e’*). Linear mixed models were constructed using patient number as random factor, and exercise, elevated exPAWP, and their interaction as fixed factors. For each DST parameter with potential to identify elevated exPAWP, a receiver operating characteristic curve was determined, and area under the curve (AUC) was calculated with the trapezoidal rule. 95% confidence intervals (CI) were calculated using stratified bootstrap replicates. AUC were compared using Delong’s test.

Holm method was used as correction for multiple comparisons. A two-sided p-value <0.05 was considered significant. All data was analyzed using R v3.6.3 (R Foundation for Statistical Computing) with packages *FSA, irr, multcomp, nlme, pROC, plotROC, rcompanion*, and *tidyverse*.

## Results

### Population

A total of 398 patients met inclusion criteria, of which 22 patients underwent exRHC, and 376 patients underwent only non-invasive DST. Compared to the exRHC cohort, patients in the DST cohort had a lower prevalence of coronary artery disease, but otherwise similar baseline characteristics (Table 1).

**Table 1:** Baseline characteristics of the study populations.

| Characteristic | ExRHC cohort<br>(n=22) | DST cohort<br>(n=376) | P<br>value |
| --- | --- | --- | --- |
| Age, years | 65 (57-71) | 68 (60-74) | .138 |
| Female sex | 10 (45) | 221 (59) | .313 |
| Heart rate, bpm | 68 (62-72) | 70 (63-82) | .435 |
| Systolic blood pressure, mmHg | 150 (127-155) | 140 (124-154) | .442 |
| BMI, kg/m<sup>2</sup> | 27.5 (26.1-30.9) | 27.8 (24.7-31.2) | .929 |
| **Past medical history** |  |  |  |
| Atrial fibrillation | 5 (23) | 78 (21) | .999 |
| Coronary heart disease | 8 (36) | 57 (15) | **.022** |
| Diabetes | 3 (14) | 51 (14) | .999 |
| Hypertension | 11 (50) | 212 (56) | .684 |
| Valvular heart disease | 1 (5) | 9 (2) | .999 |
| **Medication use** |  |  |  |
| ACE inhibitor or ARB | 6 (27) | 93 (25) | .999 |
| Aldosterone antagonist | 4 (18) | 40 (11) | .483 |
| Beta blocker | 11 (50) | 108 (29) | .064 |
| Calcium antagonist | 4 (18) | 41 (11) | .536 |
| Diuretic | 5 (23) | 74 (20) | .999 |
| Nitrate | 2 (9) | 12 (3) | .416 |
| **Laboratory analysis** |  |  |  |
| Hemoglobin, g/dL | 13.8 (12.7-15.0) | 13.8 (12.8-14.7)<br>(n=243) | .577 |
| EGFR, mg/dL | 72 (67-80)<br>(n=16) | 73 (58-86)<br>(n=218) | .481 |

Continuous variables: median (IQR), P value from Mann-Whitney-U test. Categorical variables: no. (%), P value from Chi-square test. ACE = angiotensin conversion enzyme, ARB = angiotensin receptor blocker, BMI = Body mass index, EGFR = Estimated glomerular filtration rate using CKD-EPI formula.

### Derivation of peak exercise S’ as surrogate for elevated cardiac filling pressures

In the exRHC cohort, PAWP ≥25mmHg during exercise was recorded in 14 patients, while 8 patients had normal exPAWP (Supplemental Figure 2). Comparison of baseline characteristics revealed older age, lower heart rate, more beta blocker use, and worse renal function in patients with elevated exPAWP (all p<0.05, Supplemental Table 1).

Among echocardiographic parameters, peak exercise septal systolic velocity on color Doppler (*exS’*), *exE/e’*, peak sPAP, mPAP/CO slope, peak cardiac index, and rest LV mass index were associated with elevated exPAWP (Figure 1, Supplemental Table 2). Invasive and CPET parameters that were associated with exPAWP ≥25 mmHg are displayed in Supplemental Table 2.

*ExS’* was the best echocardiographic parameter associated with elevated exPAWP, with an AUC of 0.97 (CI 0.92-1.0), compared to 0.88 (CI 0.72-1.0) for mPAP/CO slope, 0.79 (CI 0.58-0.99) for peak cardiac index, 0.76 (CI 0.55-0.96) for *exE/e’*, and 0.76 (CI 0.54-0.97) for peak sPAP (Figure 2, Supplemental Figure 3). *ExS’* had a significantly higher AUC compared to *exE/e’* (p=0.039) and peak sPAP (p=0.035), but not to mPAP/CO slope (p=0.239) or peak cardiac index (p=0.099).

**Figure 2:**
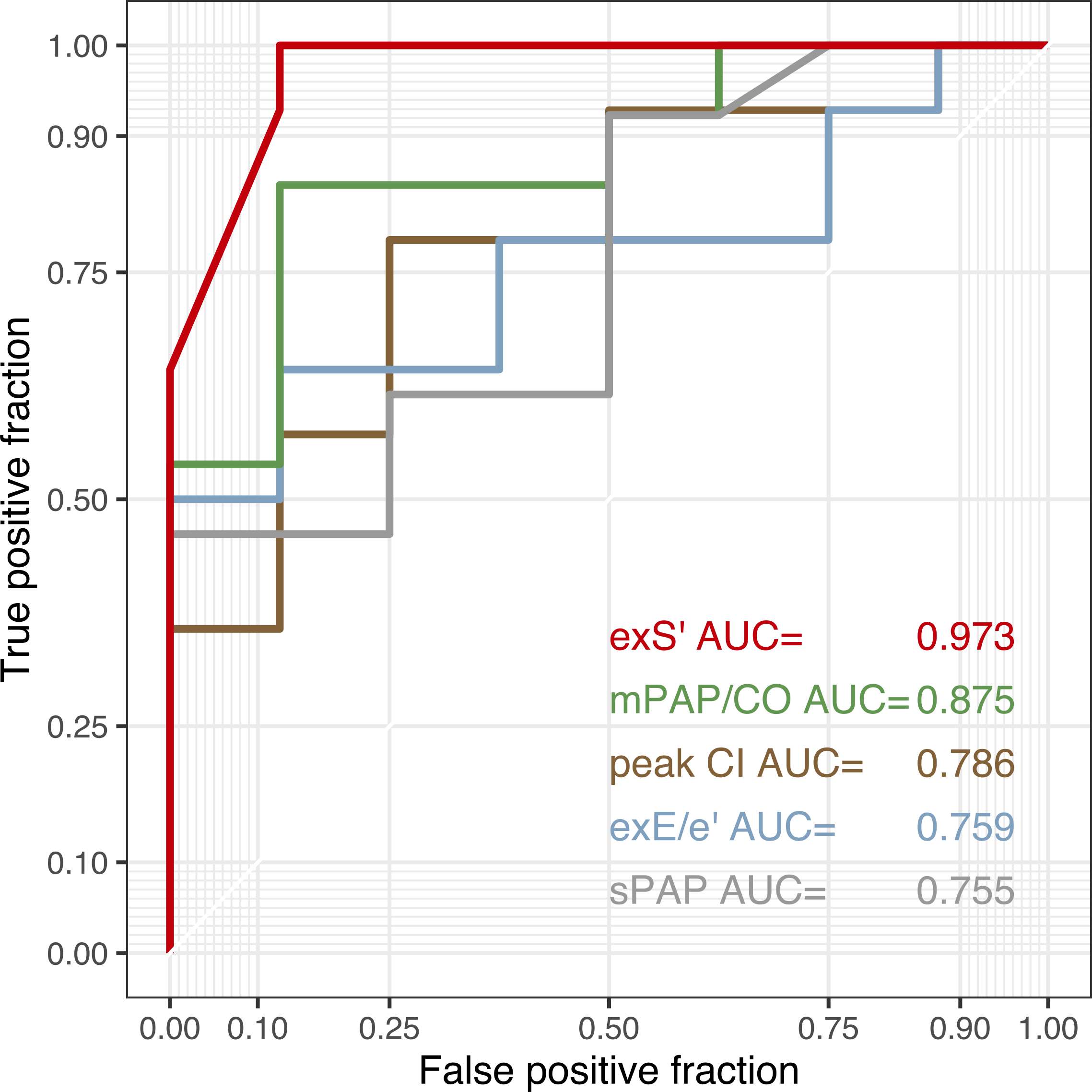
Receiver operating characteristic curves. Receiver operating characteristic curves for identifying elevated exPAWP using *exS*’ (red), mPAP/CO slope (green), peak CI (brown), *exE/e*’ (blue) and peak sPAP (grey). AUC = area under the receiver operating characteristic curve, CI = cardiac index, CO = cardiac output, exE/e’ = highest E/e’ recorded during exercise, exS’ = S’ at peak exercise, mPAP = mean pulmonary artery pressure, sPAP = systolic pulmonary artery pressure.

A threshold of *exS*’<9.5 cm/s had a specificity of 88% and sensitivity of 100% for detecting exPAWP ≥25 mmHg. *ExE/e’* ≥15 had a specificity of 100% and sensitivity of 50%; mPAP/CO slope ≥3.2 mmHg/L had a specificity of 63% and sensitivity of 85%.

As a sensitivity analysis, elevated cardiac filling pressures were alternatively defined as PAWP/CO slope >2.0 mmHg/L. AUC were comparable to the standard definition for *exS’* (0.94, CI 0.84-1.0), *exE/e’* (0.85, CI 0.68-1.0), peak sPAP (0.72, CI 0.46-0.98), and mPAP/CO slope (0.72, CI 0.46-0.97), but lower for peak cardiac index (0.44, CI 0.17-0.70).

### Decision tree for determining probability of HFpEF in inconclusive DST

In the exRHC cohort, 6/22 patients had a positive DST according to current recommendations (*exE/e’* ≥15, exTR >2.8 m/s and baseline *e*’ <7 cm/s) ^9,10^. All these patients indeed had exPAWP ≥25 mmHg. In one patient, HFpEF could be excluded according to DST recommendations, and this patient indeed had normal exPAWP. Thus, 8 patients remained with elevated exPAWP and inconclusive DSE. However, all 14 patients with elevated exPAWP had *exS*’ <9.5 cm/s. A decision tree consisting of guideline recommendations in a first step and low *exS*’ in a second step (Figure 3A), would successfully identify all patients with elevated exPAWP, at the cost of 1 false positive patient (exPAWP = 23 mmHg).

**Figure 3:**
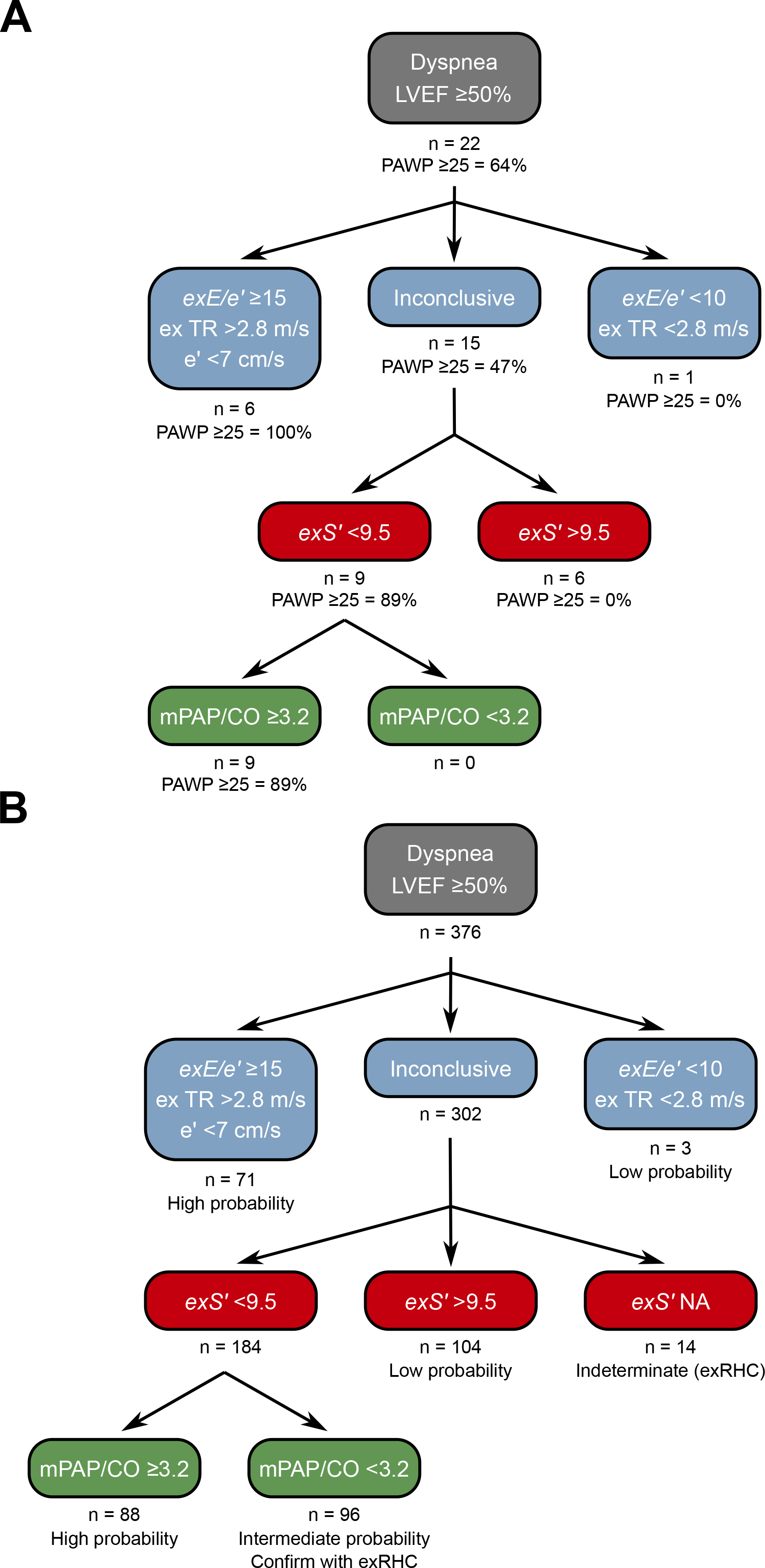
Proposed decision tree for diagnosis of HFpEF on DST. **A:** Derivation of the decision tree in the exRHC cohort. Step 1: the existing approach in ASE/EAVCI recommendations is maintained. Step 2: *exS*’ is determined and HFpEF is confirmed if <9.5 cm/s. Thus all patients with exPAPW ≥25 mmHg are identified. A single patient is false positive using this approach. Adding mPAP/CO does not provide additional information in the exRHC cohort. **B:** Application of the decision tree to the non-invasive DST cohort. Of patients with a positive DST according to ASE/EACVI recommendations, 98% had *exS’* <9.5 cm/s. None of the patients with ‘normal’ DST according to ASE/EACVI recommendations had *exS’* <9.5 cm/s. Of 302 patients with inconclusive results according to ASE/EACVI recommendations, 104 (34%) had *exS’* >9.5 cm/s, we propose that probability of HFpEF is low in these patients. A total of 184 patients (61% of inconclusive results) had *exS*’ <9.5 cm/s. Of these, 88 had elevated mPAP/CO slope suggesting exercise pulmonary hypertension, we propose that probability of HFpEF is high in these patients. In the 96 patients with normal mPAP/CO slope, we propose to perform additional investigations before establishing a diagnosis of HFpEF.

Most patients with clinically relevant HFpEF exhibit pulmonary hypertension during exercise ^5,17^. Indeed all patients with *exS’*<9.5 and PAWP ≥25 mmHg had mPAP/CO slope ≥3.2 mmHg/L. Moreover, mPAP/CO slope was the second best parameter in the AUC analysis. Thus, we suggest an algorithm based on a first step assessing *exE/e’*, adding *exS*’ in a second step, and mPAP/CO slope in a third step (Figure 3A-B).

### Applying the decision tree in the DST cohort

In the DST cohort, using ASE/EACVI recommendations a diagnosis of HFpEF was made in 71 out of 376 patients (19%). HFpEF was excluded on DST in 3 patients (1%). A total of 302 patients (80%) with inconclusive results remained.

Figure 3B shows the application of the proposed decision tree in the DST cohort. Of the 302 patients with inconclusive DST, 184 patients (61%) had abnormal *exS*’. Of the 3 patients in which HFpEF was considered unlikely based on guideline recommendations, none had abnormal *exS*’. Most of the 71 patients in which HFpEF was diagnosed based on ASE/EACVI recommendations (68 patients, 96%) had abnormal *exS*’.

Applying the proposed decision tree reduced the number of inconclusive tests from 302 (80%) to 110 (29%). A total of 192 patients (64% of inconclusive tests) could be reclassified as ‘high probability of HFpEF’ or ‘low probability of HFpEF’. Patients in the ‘high probability’ group had a worse exercise capacity compared to patients with intermediate or low probability: lower peak VO_2_ (Figure 4A), lower peak heart rate, lower workload, and steeper VE/VCO_2_ slope (Table 2). Patients classified as ‘high probability’ had a higher logistic H2FPEF score compared to patients with intermediate or low probability, indicating high likelihood of elevated exPAWP (Figure 4B, Table 2). Patients in the ‘high probability’ group were older, more frequently had atrial fibrillation, and had worse renal function compared to patients with intermediate or low probability (Table 2). Finally, compared to the other groups, patients classified as ‘high probability’ had higher resting *E/e*’, higher *exE/e’* and exercise sPAP, and reduced peak cardiac index (Table 2).

**Table 2:**
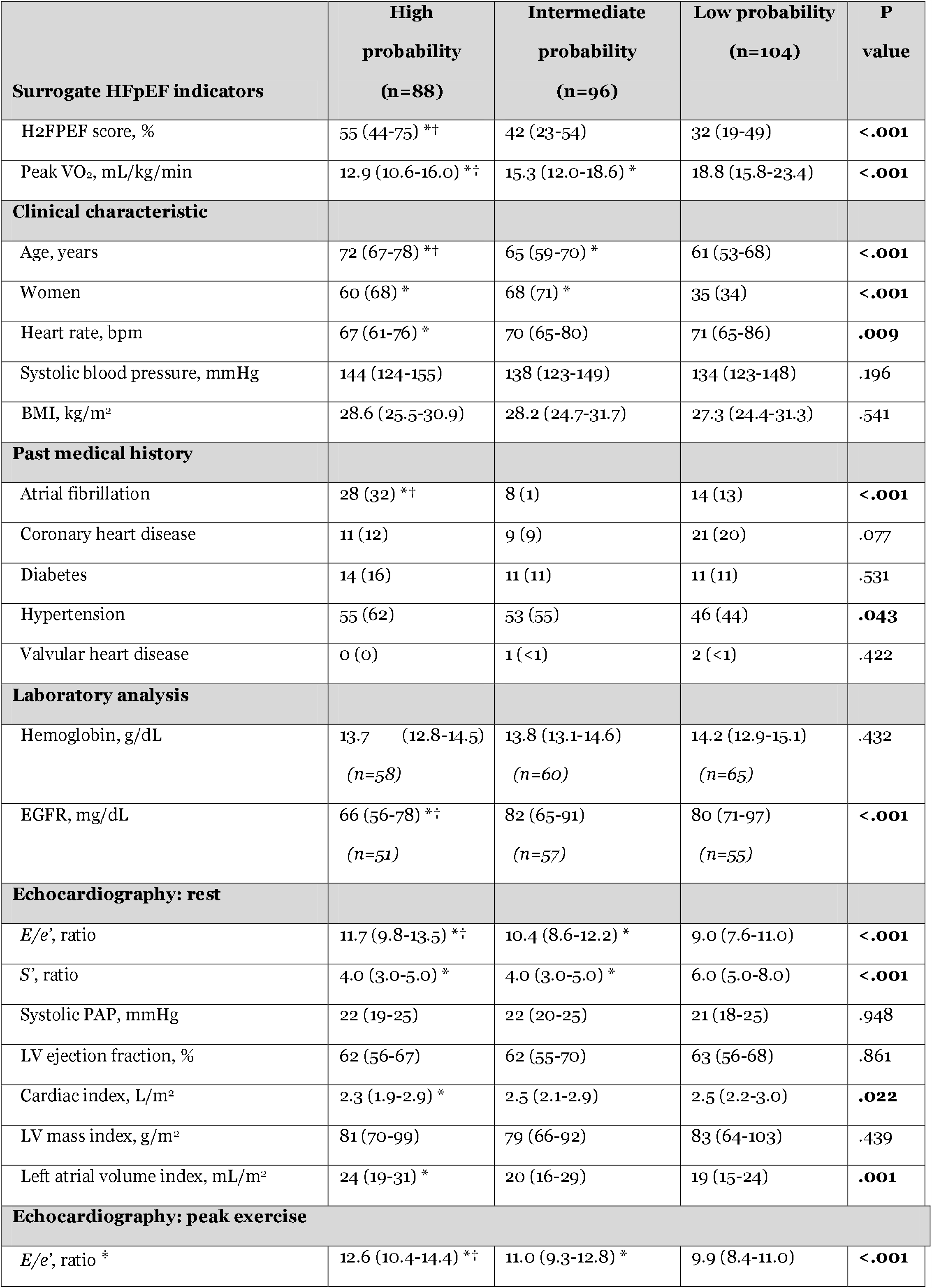
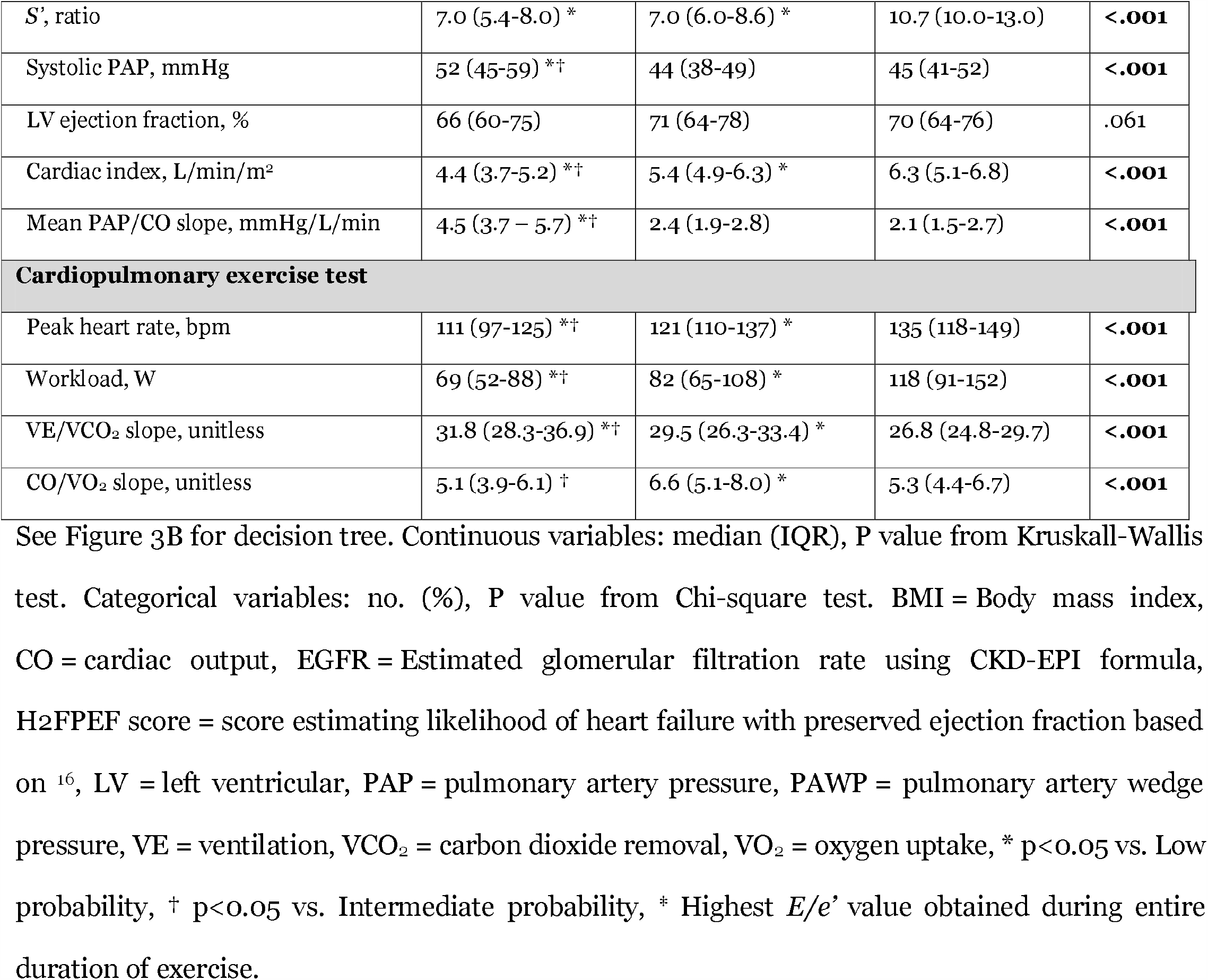
Selected characteristics and measurements in patients with inconclusive DST, stratified according HFpEF probability (decision tree).

**Figure 4:**
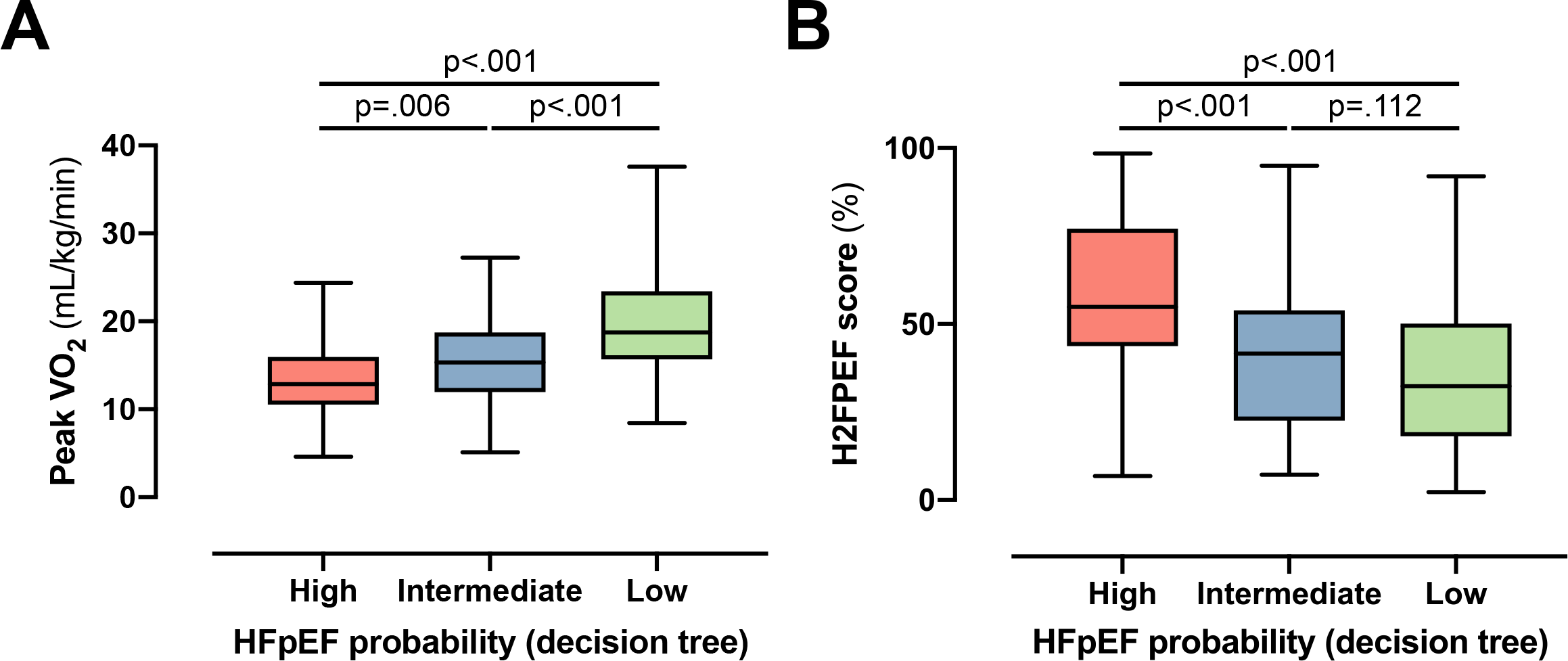
Performance of decision tree in the DST cohort. To evaluate the performance of the decision tree (Figure 3B), the probability of HFpEF according to the decision tree (high/intermediate/low) was compared to surrogate HFpEF indicators. **A:** Peak VO_2_ significantly differs between patients with high, intermediate and low probability of HFpEF according to the decision tree **B:** Logistic H2FPEF score, which calculates the probability of elevated exPAWP in percentage through clinical and echocardiographic parameters ^16^, significantly differed between patients at high and intermediate or low probability of HFpEF according to the decision tree. Multiplicity-corrected *P* values from Kruskal-Wallis test. HFpEF = heart failure with preserved ejection fraction, Peak VO_2_ = peak oxygen uptake.

Figure 5 shows the percentage of true and false positive tests using different DST criteria for diagnosis of HFpEF. All current criteria show a lack of sensitivity: of patients with invasively proven HFpEF, ASE/EACVI recommendations detected 43% ^9,10^, the Heart Failure Association consensus on HFpEF 21% ^7^, and *exE/e*’ alone 50%. Our decision tree detected 100% of HFpEF patients, at the cost of 13% false positives (compared to 0% for all recommendations).

**Figure 5:**
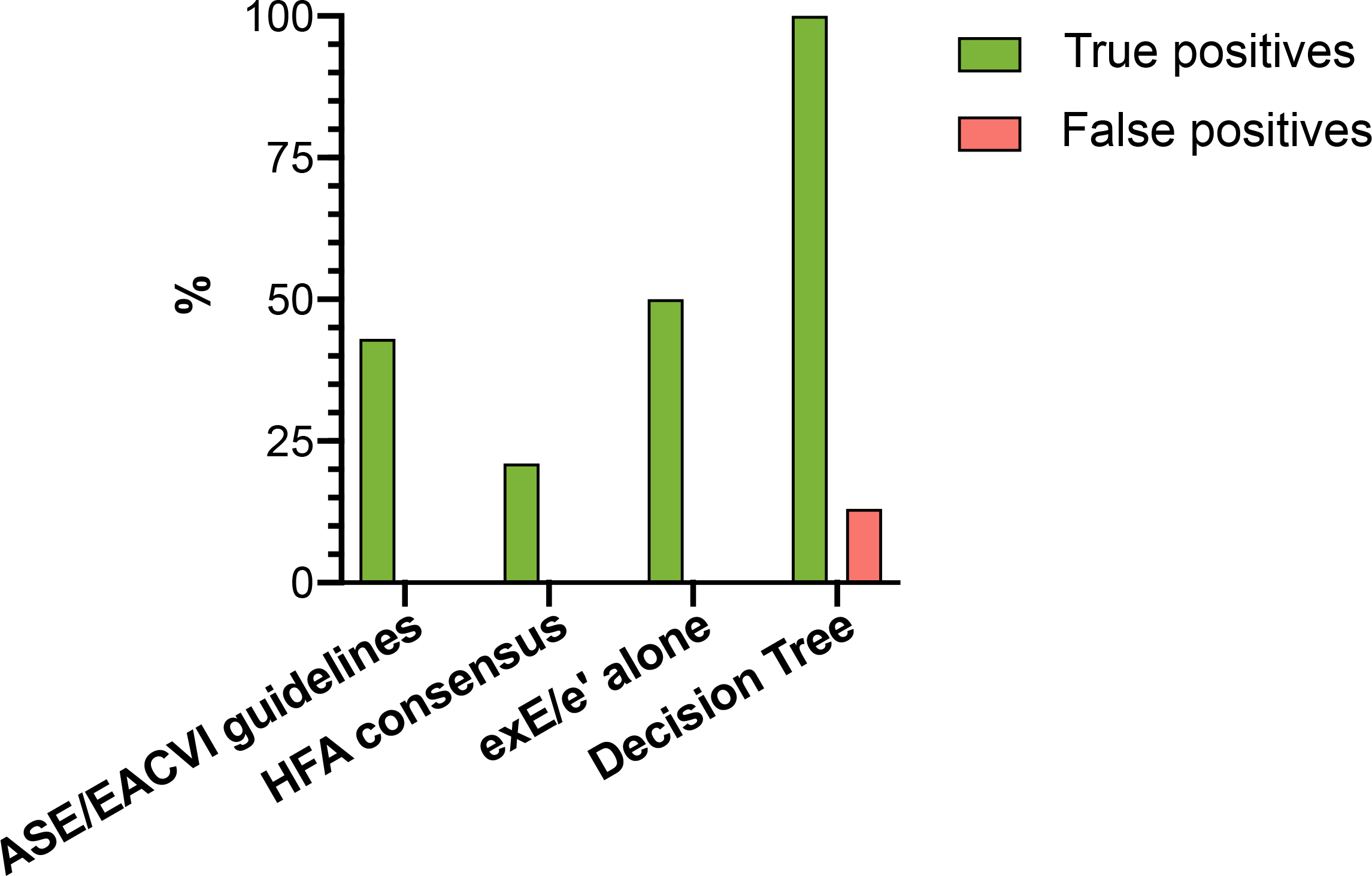
Performance of different DST algorithms for detecting elevated exPAWP. True positives: correct diagnosis of HFpEF in patients with elevated exPAWP. False positives: wrong diagnosis of HFpEF in patients with normal exPAWP. Our proposed decision tree (Figure 3) noninvasively detects 100% of true positives at the cost of 13% false positives. ASE/EACVI recommendations propose to diagnose HFpEF if *exE/e*’ is ≥15, baseline *e’* is <7 cm/s, and exercise TR is >2.8 m/s ^9,10^. The Heart Failure Association consensus (HFA-PEFF score) proposes stricter thresholds: exercise TR >3.4 m/s and average (not septal) *exE/e*’ ≥15 ^7^. *ExE/e*’ alone considers *exE/e*’ ≥15. These approaches all detect ≤50% of true positives.

### Reproducibility of peak exercise S’

*ExS’* was measured successfully in all patients in the exRHC cohort, and in 362 patients (96%) in the DST cohort. *ExS’* measured by color TDI was highly reproducible, with an interobserver agreement of 0.97 (CI 0.92-0.99). Measurement of mPAP/CO showed good interobserver agreement of 0.73 (CI 0.53-0.87). Bland-Altman plots are provided in Supplemental Figure 4.

## Discussion

In this study, we established septal *exS*’ as a compelling parameter to improve identification of elevated cardiac filling pressures in a small cohort of patients referred for simultaneous exRHC and DST. A threshold of *exS*’ <9.5 cm/s had a high sensitivity and specificity to identify exPAPW ≥25 mmHg. We propose a decision tree to diagnose HFpEF on DST, incorporating *exS’* and mPAP/CO slope. Applying this decision tree to 376 patients with suspected HFpEF substantially improved the diagnostic yield of DST from 20% (using guideline recommendations) to 71% (using the decision tree).

Current ASE/EACVI recommendations recommend the use of *exE/e’* and sPAP to diagnose HFpEF on DST ^9,10^. These recommendations are based on early studies focusing solely on *exE/e’*, disregarding other possible correlates of elevated exPAWP ^18^. Most of these were performed without concurrent exPAWP measurement, and subsequent invasive validation studies showed at most a moderate correlation between *exE/e’* and PAPW ^11,19^. Another limitation of the evaluation of *exE/e*’ relates to the influences of increased respiratory rate and tachycardia that occur during exercise. Hence, fusion of E/A waves and e’/a’ waves often occurs beyond heart rates of 100 bpm, thereby compromising the accuracy of this assessment. *ExE/e’* has a good specificity for diagnosis of elevated exPAWP, but shows poor sensitivity ^11,18^. This leaves many DST with an inconclusive result, up to 80% in our population.

A recent Heart Failure Association expert consensus paper proposed DST in patients with an intermediate to high pre-test probability of HFpEF ^7^. However, the authors suggested a stricter cutoff of >3.4 m/s for exercise TR, which further reduces sensitivity, as confirmed by our study findings.

It is well accepted that patients with HFpEF not only have impaired diastolic cardiac function, but also suffer from subtle reductions in systolic function despite a normal LVEF ^20^. Measurements of longitudinal function, such as strain and strain rate, have emerged as less afterload dependent surrogates of systolic function, but are affected by respiratory variation in image quality at peak exercise. In contrast, systolic velocity of the mitral annulus (*S’*) can be easily obtained at peak exercise regardless of heart rate and image quality (in 96% of patients in our study), while showing high reproducibility. From a mechanistic point of view, the reduction of *exS*’ in patients with increased exPAWP during exercise may be explained by decreased diastolic suction and elastic recoil resulting from a lack of systolic functional reserve. Hence, as the capacity of the LV to decrease its end-systolic volume during exercise is reduced, the driving force for early diastolic suction to enable is impaired and rapid LV filling becomes exquisitely dependent on increased filling pressures across the mitral valve.

Other studies have previously evaluated longitudinal LV function during exercise in HFpEF patients. Wang *et al*. found reduced values of resting *S*’ and *exS*’ in HFpEF patients compared to controls ^21,22^. *ExS’* correlated well with peak VO_2_ ^23,24^, and was a significant predictor of all-cause mortality and HF hospitalization ^25^. Also, right ventricular S’ was demonstrated to be a useful marker of longitudinal function during exercise, and was able to identify athletes at risk of arrythmias ^26^.

Using our proposed decision tree (Figure 3B), 64% of patients with inconclusive DST could be reclassified as high or low probability of HFpEF, substantially improving the diagnostic yield of DST. In the decision tree, we maintain *exE/e’* in the first step because of its extensive validation in multiple populations, and its high specificity ^11,18,27^. Note that we use a threshold of ≥15 because we routinely record only septal e’, when using average *E/e’* a threshold of ≥14 is more appropriate ^9^. In a next step, *exS’* is evaluated and HFpEF is confirmed for a value <9.5 cm/s based on our current findings. As the group of patients with low *exS’* remains large, we decided to add evidence of exercise pulmonary hypertension as a third step. In our opinion, an indication of exercise pulmonary hypertension must be present for the diagnosis of HFpEF using DST alone, because of the close pathophysiological relation between left atrial pressure, PAWP and mPAP. We chose mPAP/CO slope rather than sPAP, because (1) mPAP/CO slope is more accurate in situations where peak exercise CO is abnormal, such as in HFpEF ^14^, (2) mPAP/CO was the next-to-best parameter in the AUC analysis, and (3) in the exRHC cohort all patients with *exS*’<9.5 cm/s and elevated exPAWP had a mPAP/CO slope above threshold.

In a number of patients, exercise pulmonary hypertension was not present, but *exS’* <9.5 cm/s indicated elevated exPAWP. This may reflect early HFpEF in patients with relatively compliant left atrium and pulmonary vasculature, underestimation of mPAP/CO slope on DST, or lower specificity of *exS*’ in an unselected population. In these cases, other methods can aid to establish a final diagnosis of HFpEF. The gold standard investigation for these patients remains an exRHC, as sPAP and mPAP/CO slope are generally underestimated on echocardiography when compared to invasive measurement ^14^.

Our study results should be interpreted in the context of some limitations. Color TDI is angle-dependent, however the use of offline repositioning and the use of septal rather than lateral *S’* mitigated the impact of this limitation. Whether using a pulse wave TDI signal optimized for assessing *S’* has equal diagnostic capabilities, remains to be studied.

A ‘gold standard’ to diagnose HFpEF non-invasively is currently still lacking. As such, we used several surrogate measures (peak VO_2_, logistic H2FPEF score) and supporting features (diastolic function, typical clinical characteristics) in the DST cohort to demonstrate differences between patients classified as high, intermediate or low probability of HFpEF.

Furthermore, the small sample size of the exRHC cohort compared to the DST cohort suggests a highly selected population. Our results should be validated in a larger patient cohort.

We conclude that *exS’* was the most accurate parameter to identify patients with elevated cardiac filling pressures in a cohort of patients referred for exRHC because of exertional dyspnea. We propose a decision tree to diagnose elevated exPAWP on DST in patients with exertional dyspnea and LVEF ≥50%. Applying this decision tree for the diagnosis of HFpEF on DST substantially improved the diagnostic yield from 20% (using guideline recommendations alone) to 71% (using the decision tree). Validation in a separate exRHC cohort is desirable before application of our findings in clinical practice.

## Data Availability

The data is accessible for other researchers by request to the authors.

## Acknowledgements

We thank the sonographers, nurses and cardiology fellows at the Department of Cardiology, Jessa Hospital, Hasselt for their invaluable assistance during the exRHC and DST procedures.

