## Supplemental Data for "Novel echocardiographic markers of elevated left ventricular filling pressure during diastolic stress testing"

Supplemental Figure 1: Representative example of color tissue Doppler image for measuring peak exercise *S’*

Supplemental Figure 2: PAWP evolution during exercise in the exRHC cohort

Supplemental Figure 3: Relation between exPAWP and *exS’* in the exRHC cohort

Supplemental Figure 4: Bland-Altman plots of the novel echocardiographic parameters.

Supplemental Table 1: Baseline characteristics of the exRHC cohort, stratified according to PAWP ≥ or < 25 mmHg

Supplemental Table 2: Echocardiography, invasive hemodynamics, arterial blood gas measurement and cardiopulmonary exercise test results in the exRHC cohort

Supplemental File 1 (Supplemental_File_1.mp4): Representative example of color tissue Doppler image at peak exercise.

**Supplemental Figure 1: Representative example of color tissue Doppler image for measuring peak exercise *S’*.** Peak exercise *S’* was 8 cm/s in this patient. This was measured at the 4^th^ cardiac cycle in this recording (red line). Note that the first steep upstroke in the tissue Doppler signal (red arrow) should NOT be measured. Also note the small variability of *S’* compared to the large variability in *e’* at peak exercise.

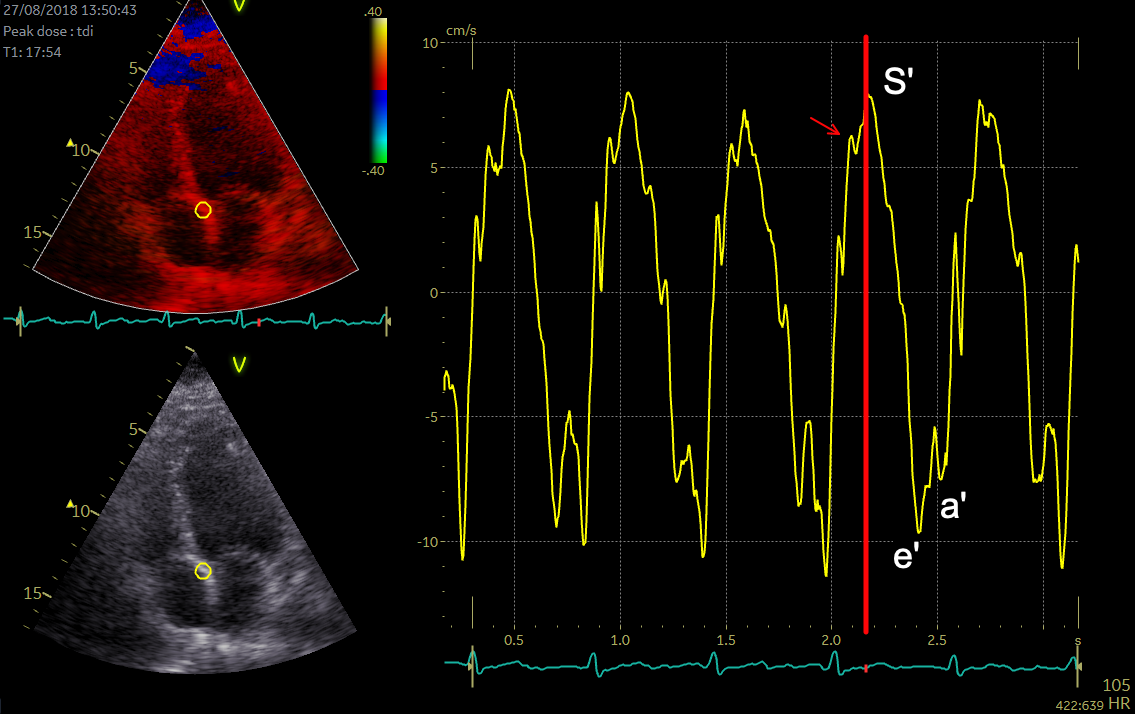

**Supplemental Figure 2: PAWP evolution during exercise in the exRHC cohort.** PAWP was measured at rest, at 3, 6 and 9 minutes of exercise, and at peak exercise. Red dots: patients with PAWP ≥25 mmHg at peak exercise (n=14), green dots: patients with PAWP <25 mmHg at peak exercise (n=8), lines: linear regression. Mean with standard error. exPAWP = pulmonary artery wedge pressure at peak exercise, exRHC = exercise right heart catheterization, PAWP = pulmonary artery wedge pressure.

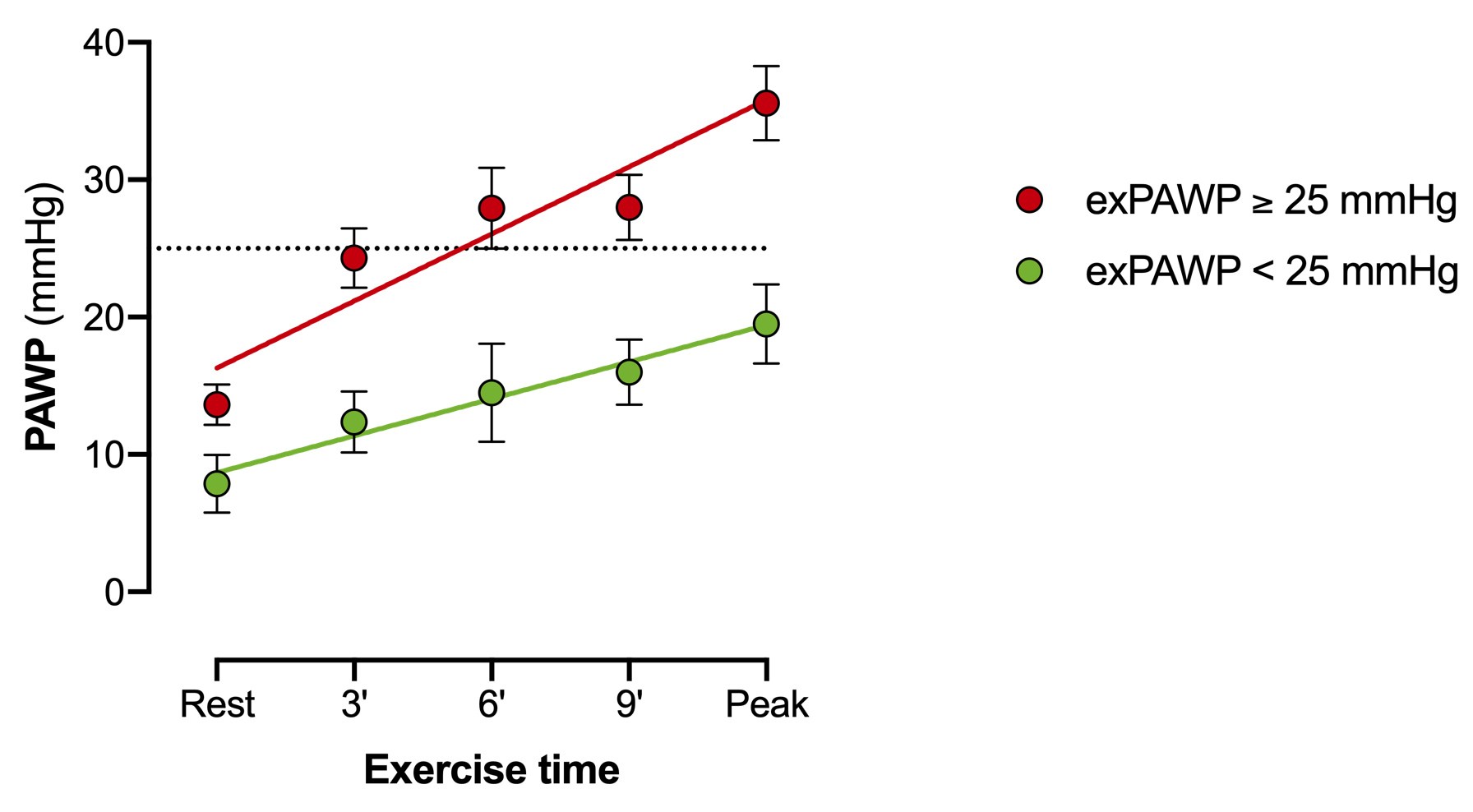

**Supplemental Figure 3: Relation between exPAWP and *exS’* in the exRHC cohort.** Dashed lines: thresholds for elevated exPAWP (≥25 mmHg) and low *exS’* (<9.5 cm/s). Upper left quadrant: true positives (elevated exPAWP and low *exS’*), lower left quadrant: false positives (low exPAWP and low *exS’*), lower right quadrant: true negatives (low exPAWP and high *exS’*). exPAWP = pulmonary artery wedge pressure at peak exercise, exRHC = exercise right heart catheterization, *exS’* = peak systolic annular velocity.

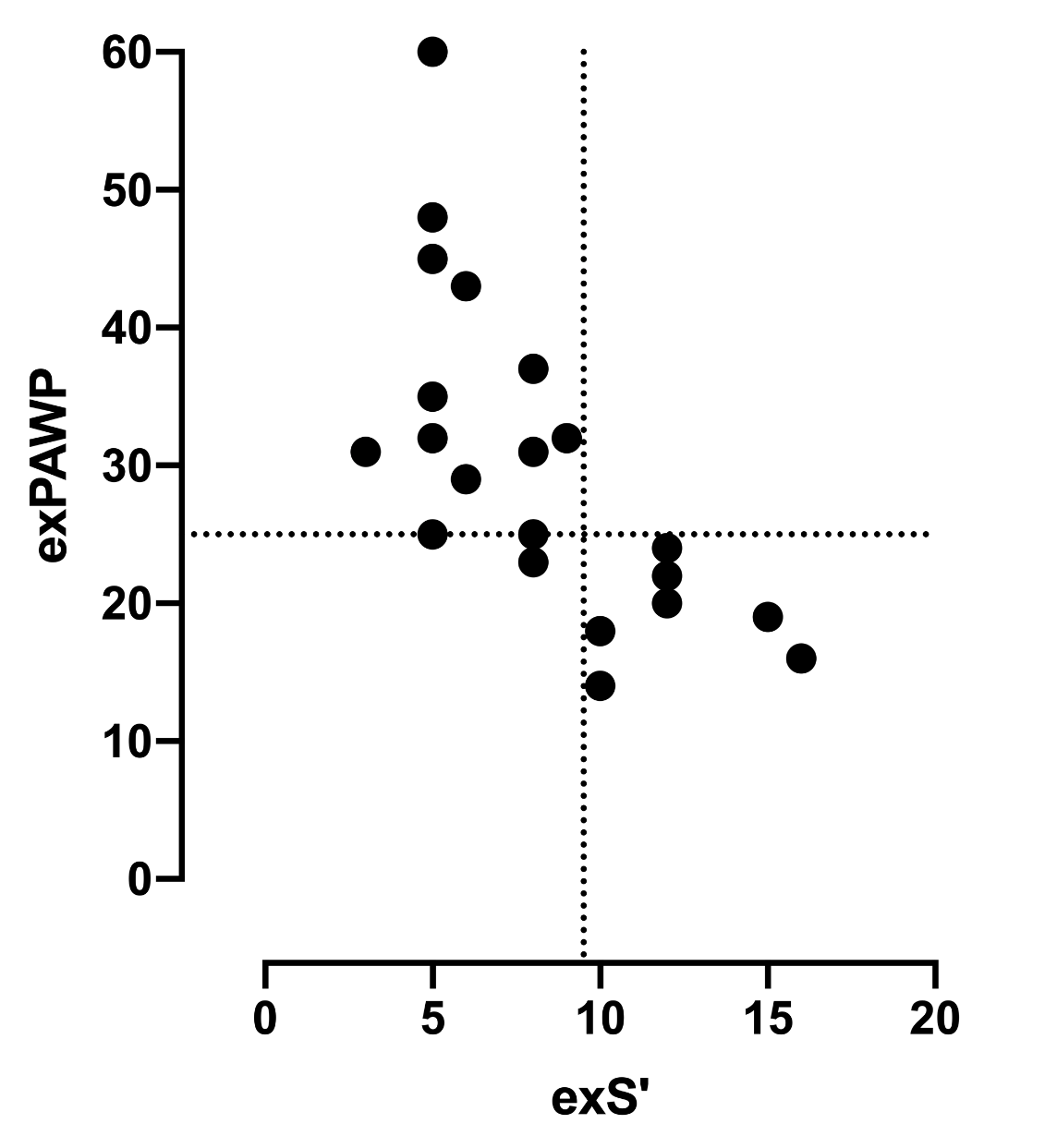

**Supplemental Figure 4: Bland-Altman plots of key echocardiographic parameters.** Difference vs average plot of key echocardiographic parameters for 22 patients of the DST cohort and 3 observers. **A: $** *exS’* as cm/s. **B:** mPAP/CO as mmHg•L^-1^•min^-1^ **C**: peak mPAP as mmHg, calculated by the Chemla formula as sPAP*0.61+2 with sPAP estimated from colloid-enhanced tricuspid regurgitation signal without adding right atrial pressure, **D:** peak CO as L•min^-1^, calculated by the left ventricular outflow tract method (**D**). Dashed lines: upper and lower limits of agreement, full line: mean difference (bias). CO = cardiac output, *exS’* = peak exercise septal systolic velocity on color Doppler, mPAP = mean pulmonary artery pressure, SD = standard deviation, sPAP = systolic pulmonary artery pressure.

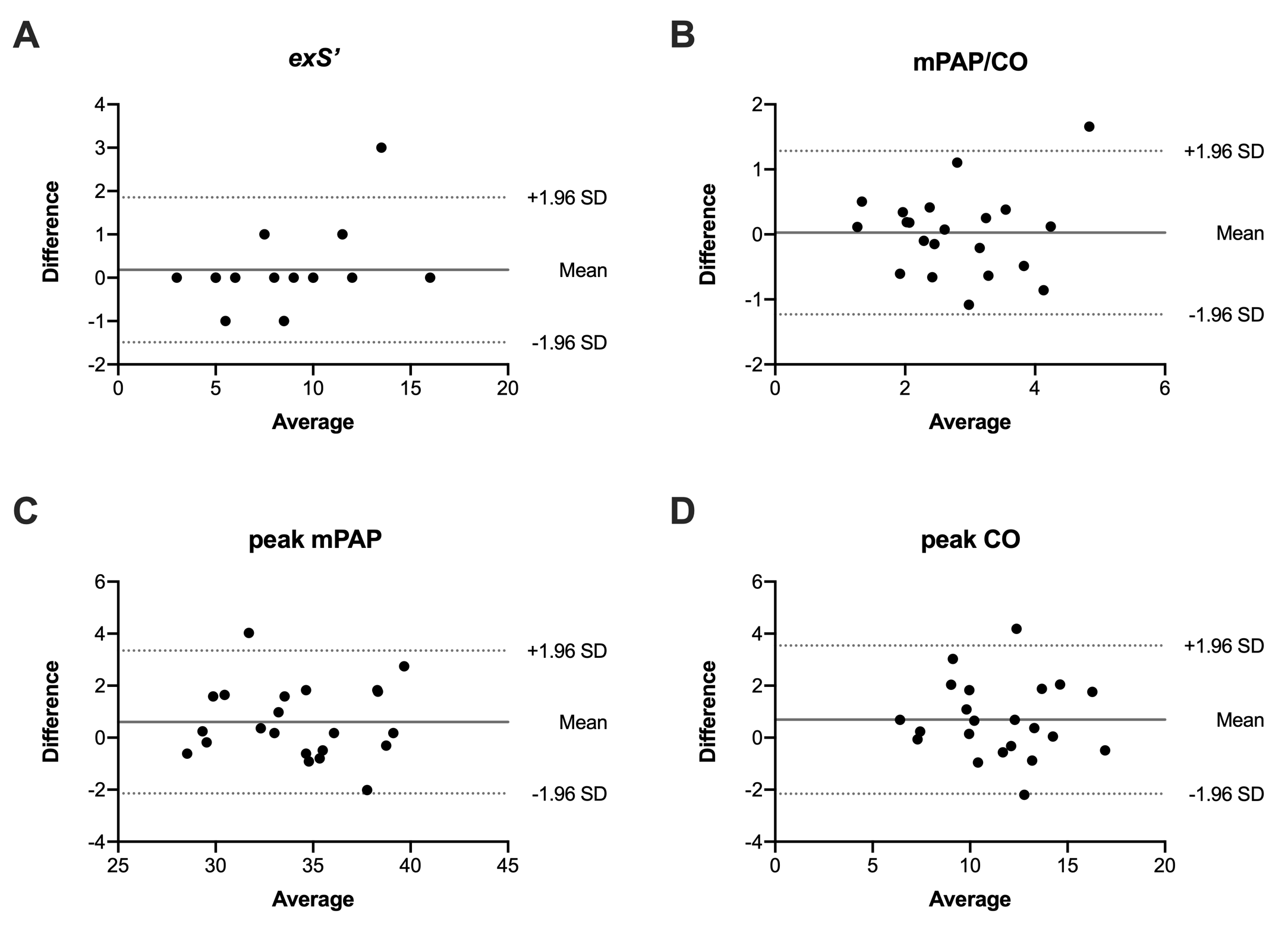

**Supplemental Table 1: Baseline characteristics of the exRHC cohort, stratified according to PAWP ≥ or < 25 mmHg**

| **Characteristic** | **PAWP  ≥25 mmHg (n=14)** | **PAWP  <25 mmHg (n=8)** | **P value** |
| --- | --- | --- | --- |
| Age, median (IQR), years | 67 (64-73) | 59 (55-62) | **.027** |
| Female sex, no. (%) | 7 (50) | 3 (38) | .675 |
| Heart rate, median (IQR), bpm | 66 (59-71) | 72 (70-78) | **.044** |
| Systolic blood pressure, median (IQR), mmHg | 151 (137-156) | 128 (127-151) | .673 |
| BMI, median (IQR), kg/m^2^ | 28.2 (26.2-31.8) | 26.7 (25.0-28.1) | .238 |
| **Past medical history** |  |  |  |
| Atrial fibrillation, no. (%) | 4 (29) | 1 (13) | .736 |
| Coronary heart disease, no. (%) | 8 (57) | 0 (0) | **.018** |
| Diabetes, no. (%) | 2 (14) | 1 (13) | .999 |
| Hypertension, no. (%) | 9 (64) | 2 (25) | .183 |
| Valvular heart disease, no. (%) | 1 (7) | 0 (0) | .999 |
| **Medication use** |  |  |  |
| ACE inhibitor or ARB, no. (%) | 6 (42) | 0 (0) | .051 |
| Aldosterone antagonist, no. (%) | 4 (29) | 0 (0) | .254 |
| Beta blocker, no. (%) | 10 (71) | 1 (13) | **.024** |
| Calcium antagonist, no. (%) | 3 (21) | 1 (13) | .999 |
| Diuretic, no. (%) | 4 (29) | 1 (13) | .613 |
| Nitrate, no. (%) | 2 (14) | 0 (0) | .515 |
| **Laboratory analysis** |  |  |  |
| Hemoglobin, median (IQR), g/dL | 13.6 (13.3-14.4) | 15.0 (14.6-16.2) | .147 |
| EGFR, median (IQR), mg/dL | 68 (65-72)  *(n=8)* | 75 (72-102) | **.021** |

P value from Mann-Whitney-U test (continuous variables) or Fisher’s exact test (categorical variables). ACE = angiotensin conversion enzyme, ARB = angiotensin receptor blocker, BMI = Body mass index, EGFR = Estimated glomerular filtration rate using CKD-EPI formula.

**Supplemental Table 2: Echocardiography, invasive hemodynamics, arterial blood gas measurement and cardiopulmonary exercise test results in the exRHC cohort**

| **Echocardiography** | **Elevated exPAWP** | | **Normal exPAWP** | | **P-Group** | **P-Exercise** | **P-Inter-action** | **P Peak vs. Peak** |
| --- | --- | --- | --- | --- | --- | --- | --- | --- |
|  | **Rest** | **Peak** | **Rest** | **Peak** |  |  |  |  |
| *E/e’*,  median (IQR) * | 11.1  (9.0-15.9) | 14.7  (11.9-18.4) | 8.4  (7.4-9.6) | 11.1  (9.0-12.9) | .090 | .049 | .798 | **.043** |
| *S’,*  median (IQR), cm/s | 4.0  (3.1-4.8) | 5.5  (5.0-8.0) | 6.0  (5.0-7.2) | 12.0  (10.0-12.8) | .025 | .002 | .002 | **<.001** |
| Systolic PAP,  median (IQR), mmHg | 21  (18-24) | 59  (45-69) | 20  (18-22) | 43  (32-56) | .834 | <.001 | .026 | **.005** |
| LV ejection fraction,  median (IQR), % | 66  (59-70) | 74  (58-78) | 62  (60-64) | 70  (60-72) | .571 | .049 | .857 | .999 |
| LV end diastolic volume index,  median (IQR), mL/m^2^ | 48  (40-55) | 46  (41-50) | 44  (41-52) | 39  (36-46) | .516 | .212 | .623 | / |
| Cardiac index,  median (IQR), L/min/m^2^ | 2.3  (2.2-3.6) | 5.1  (4.4-5.8) | 2.6  (2.5-2.9) | 6.1  (5.8-6.5) | .884 | <.001 | .017 | **.026** |
| RV fractional area change,  median (IQR), % | 54  (50-59) | 56  (52-65) | 50  (49-58) | 55  (53-63) | .778 | .180 | .929 | / |
| LV mass index,  median (IQR), g/m^2^ | 90  (75-116) | | 62  (55-74) | | **.006** | / | / | / |
| Left atrial volume index,  median (IQR), mL/m^2^ | 19.8  (14.9-24.6) | | 17.5  (16.7-27.2) | | .827 | / | / | / |
| Mean PAP/CO slope,  median (IQR), mmHg/L | 4.8  (3.9-5.9) | | 2.5  (1.1-3.3) | | **.003** | / | / | / |
| CO/VO_2_ slope, median (IQR) | 5.4 (4.6-6.3) | | 4.9 (3.6-6.8) | | .547 | / | / | / |
| **Invasive hemodynamics** | **Elevated exPAWP** | | **Normal exPAWP** | | **P-Group** | **P-Exercise** | **P-Inter-action** | **P Peak vs. Peak** |
|  | **Rest** | **Peak** | **Rest** | **Peak** |  |  |  |  |
| Heart rate,  median (IQR), bpm | 70  (59-83) | 121  (114-136) | 71  (69-83) | 144  (128-158) | .779 | <.001 | .022 | .086 |
| Systolic blood pressure,  median (IQR), mmHg | 168  (150-193) | 214  (196-235) | 155  (150-163) | 206  (184-236) | .476 | .001 | .837 | .964 |
| PAWP,  median (IQR), mmHg | 13  (10-19) | 32  (30-42) | 9  (7-10) | 20  (18-22) | .071 | <.001 | .007 | **<.001** |
| Systolic PAP,  median (IQR), mmHg | 32  (30-37) | 73  (66-79) | 22  (21-26) | 50  (46-57) | .139 | <.001 | .032 | **<.001** |
| Right atrial pressure,  median (IQR), mmHg | 8  (4-11) | 18  (16-25) | 4  (3-4) | 9  (9-12) | .063 | <.001 | .044 | **<.001** |
| Diastolic pressure gradient,  median (IQR), mmHg | 1  (0-2) | 5  (3-11) | 1  (1-5) | 8  (4-12) | .773 | .012 | .275 | .401 |
| Cardiac index,  median (IQR), L/min/m^2^ | 2.2  (1.9-2.5) | 5.1  (4.3-6.1) | 2.5  (2.2-3.2) | 6.5  (6.0-8.3) | .271 | <.001 | .534 | .167 |
| Pulmon. vascular resistance,  median (IQR), dynes*s/cm^5^ | 136  (102-176) | 123  (97-200) | 114  (63-129) | 107  (51-125) | .200 | .704 | .961 | / |
| Systemic vascular resistance,  median (IQR), dynes*s/cm^5^ | 2111  (1809-2640) | 907  (629-1262) | 1687  (1282-1953) | 652  (545-710) | .030 | <.001 | .289 | .648 |
| PAWP/CO slope,  median (IQR), mmHg/L | 3.4  (1.5-5.6) | | 1.2  (1.0-1.6) | | **.016** | / | / | / |
| Mean PAP/CO slope,  median (IQR), mmHg/L | 6.0  (3.7-8.5) | | 2.4  (2.0-3.3) | | **.006** | / | / | / |
| CO/VO_2_ slope, median (IQR) | 9.4 (8.2-11.5) | | 6.7 (5.3-9.2) | | .165 | / | / | / |
| **Arterial blood gas measurement** | **Elevated exPAWP** | | **Normal exPAWP** | | **P-Group** | **P-Exercise** | **P-Inter-action** | **P Peak vs. Peak** |
|  | **Rest** | **Peak** | **Rest** | **Peak** |  |  |  |  |
| Central venous O_2_ saturation,  median (IQR), % | 65  (63-67) | 38  (32-44) | 69  (69-70) | 32  (27-36) | .372 | <.001 | .060 | .198 |
| O_2_ extraction,  median (IQR), mL/dL | 5.5  (5.1-6.0) | 10.6  (8.0-12.2) | 5.2  (4.9-5.6) | 12.3  (11.5-14.0) | .599 | <.001 | .031 | .084 |
| Lactate,  median (IQR), mmol/L | 1.1  (0.9-1.3) | 5.2  (4.3-6.1) | 1.0  (0.9-1.3) | 5.7  (3.2-7.6) | .968 | <.001 | .069 | **.026** |
| **Cardiopulmonary exercise test** | **Elevated exPAWP** | | **Normal exPAWP** | | **P-Group** | **P-Exercise** | **P-Inter-action** | **P Peak vs. Peak** |
|  | **Rest** | **Peak** | **Rest** | **Peak** |  |  |  |  |
| VO_2_,  median (IQR), mL/kg/min | 3.1 (2.6-3.7) | 12.5 (10.9-14.4) | 3.7  (3.2-3.8) | 22.7  (17.8-24.0) | .610 | <.001 | .003 | **<.001** |
| Workload,  median (IQR), W | 0  (0-0) | 90  (71-111) | 0  (0-0) | 157  (109-183) | .999 | <.001 | .008 | **<.001** |
| Respiratory exchange ratio,  median (IQR) | 0.77  (0.70-0.81) | 1.13  (1.08-1.16) | 0.86  (0.82-0.86) | 1.15  (1.11-1.21) | .049 | <.001 | .611 | .338 |
| Breathing reserve,  median (IQR), % | 100  (100-100) | 47  (29-56) | 100  (100-100) | 61  (54-72) | .999 | <.001 | .776 | .664 |
| VE/VCO_2_ slope,  median (IQR) | 31.8  (28.7-35.3) | | 25.4  (23.3-26.8) | | **.013** | / | / | / |

Elevated exPAWP was defined as PAWP ≥25 mmHg during peak exericse. P values from linear mixed models analysis (2 time points) or Mann-Whitney-U test (single time point). P Peak vs Peak from Holm-corrected post hoc multiple comparisons. CO = cardiac output, exPAWP = pulmonary artery wedge pressure during peak exercise, LV = left ventricular, PAWP = pulmonary artery wedge pressure, PAP = pulmonary artery pressure, PAWP = pulmonary artery wedge pressure, VE = ventilation, VCO_2_ = carbon dioxide removal, VO_2_ = oxygen uptake, * Highest *E/e’* value obtained during exercise.

**Supplemental File 1 (Supplemental_File_1.mp4): Representative example of color tissue Doppler image at peak exercise.** Peak exercise *S’* was 8 cm/s in this patient. This was measured at the 4^th^ cardiac cycle in this recording, other cardiac cycles show a slight off-angle movement due to respiration (cycle 1, cycle 3) or slight rotation toward the left ventricular outflow tract (cycle 5). Note that the first steep upstroke should not be measured. Also note the small variability of *S’* compared to the large variability in *e’* at peak exercise.
